# Detection of Perivascular Spaces at the Gray-White Matter Interface Using Heavily T_2_-weighted MRI at 7T

**DOI:** 10.1101/2024.10.11.24315113

**Authors:** Gael Saib, Zeynep H. Demir, Paul A. Taylor, S. Lalith Talagala, Alan P. Koretsky

**Author notes:** Corresponding Author: Gael Saib, Laboratory of Functional and Molecular Imaging, National Institute of Neurological Disorders and Stroke, National Institutes of Health, 10 Center Drive, Building 10, Room B1D69, Bethesda, MD 20892, USA. These authors contributed equally to this work (Gael Saib, Zeynep H. Demir).

## Abstract

**Background:** There is increasing interest in high-contrast cerebrospinal fluid (CSF) MRI for imaging perivascular spaces (PVSs). Dilated PVSs, associated with aging, dementia, and various other conditions, are readily detected within the white matter (WM), basal ganglia, and midbrain. While 7T MRI enables detection of smaller PVSs, cortical PVS burden has received limited attention despite its potential value for understanding neurological conditions.

**Purpose:** To investigate the detectability of cortical PVS segments in healthy participants using heavily T_2_-weighted MRI at 7T.

**Materials and Methods:** A T_2_-weighted 3D-TSE sequence was optimized at 7T to detect CSF with high resolution and contrast-to-noise ratio (CNR) while minimizing signal from surrounding tissues. A semi-automated pipeline was developed to extract PVSs and quantify their density in the whole brain, including the cortex.

**Results:** Seventeen healthy volunteers (40±14 years) were scanned at 7T. Optimized TSE achieved a CSF-to-tissue CNR of ∼180:1, enabling detection of small PVSs throughout the brain and leukocortical segments. About 20% of WM PVSs contain a leukocortical segment. WM PVSs with a leukocortical segment represented 70% of the total PVS volume. PVS density in the cortex was ∼0.7% (∼6-fold lower than WM), with highest in the insula and lowest in the auditory cortex.

**Conclusion:** High-resolution CSF imaging using optimized 3D-TSE MRI at 7T allows detection and quantification of leukocortical PVS segments at the gray-white matter interface in healthy individuals. This study lays the groundwork for exploring regional PVS changes related to the cortex and their potential use in diagnosis or prognosis of neurological diseases.

## 1. Introduction

Perivascular spaces (PVSs), also known as Virchow-Robin spaces, are fluid-filled compartments surrounding blood vessels in the brain parenchyma (Durand-Fardel, 1843; Virchow, 1851). They serve as a pathway for cerebrospinal fluid (CSF) exchange with interstitial fluid (ISF) and play an important role in immune surveillance, brain homeostasis and the clearance of metabolic waste (Wardlaw et al., 2020). Emerging studies have shown that PVS dilatation is associated with aging, small vessel diseases, and neurodegenerative diseases (Francis et al., 2019). MRI has enabled the non-invasive characterization of dilated PVSs, notably along lenticulostriate arteries in the basal ganglia, medullary arteries in white matter (WM), and perforating arteries in midbrain (Kwee & Kwee, 2007). Large dilated PVSs associated with opercular and insular middle cerebral arteries (MCA) branches have also been identified in the anterior temporal lobe region and frontal operculum (McArdle et al., 2020) as well as along hippocampal perforating vessels (Yao et al., 2014). Heavily T_2_-weighted MRI at high field (>3T) has provided superior CSF-to-tissue contrast to noise ratio (CNR) and finer spatial resolution (Daoust et al., 2017; Gao et al., 2014). 7T MRI has further improved the detection of PVSs at sub-millimeter spatial resolution (Barisano et al., 2019; Bouvy et al., 2014; Zong et al., 2016), enabling earlier identification of changes in smaller PVSs.

Most arterial PVSs in the basal ganglia and centrum semiovale are coated with two layers of leptomeninges (Pollock et al., 1997), in contrast to the cortex where the periarterial space is formed by a single layer of leptomeninges. In cortical penetrating arterioles and capillaries, this layer gets increasingly compressed by glia limitans and the pia mater against the vessel wall resulting in smaller PVSs (Morris et al., 2016; E. T. Zhang et al., 1990). For subcortical arteries originating from the cortex, this transition occurs at the gray-white matter interface, defined as transition zone from cortical gray matter (GM) to WM (Song et al., 2025), and may be associated with regional differences in fluid drainage properties. As communication pathways between subarachnoid CSF and the cortex has been demonstrated (Eide et al., 2018; Ringstad et al., 2018), early changes in leukocortical PVS segments in the gray-white matter interface may offer valuable insight into neurological conditions. High-resolution MRI has enabled visualization of PVSs in this region in both healthy individuals (Liu et al., 2025) and in patients with cerebral amyloid angiopathy (Perosa et al., 2022). However, to the best of our knowledge, a detailed imaging study of cortical PVS segments has not previously been reported.

In a previous preliminary study, a high-resolution T_2_-weighted 3D Turbo Spin Echo (TSE) sequence was optimized at 7T to maximize CSF-to-tissue CNR while minimizing signal from parenchyma, with a specific attention to CSF-to-GM contrast (Saib et al., 2023). In the present work, PVSs were characterized in the whole brain, with a focus on leukocortical segments in the gray-white matter interface, using a semi-automated processing technique that accounts for local variations in signal and background noise at 7T. Based on high-resolution MP2RAGE segmentation, PVS density was quantified in the whole brain and within 266 cortical areas for leukocortical PVS segments. This study lays the groundwork for assessing the diagnostic utility of detecting PVS burden in the gray-white matter interface as well as understanding the mechanisms for structural and contrast changes in the cortex in patients with neurological disorders.

## 2. Materials and Methods

### 2.1. Study design and participants

This study was conducted under a protocol (00-N-0082) approved by the Institutional Review Board of the National Institutes of Health (Bethesda, Maryland, USA) and registered on ClinicalTrials.gov (NCT 00004577). A total of 17 healthy volunteers (10 males and 7 females; mean age 40±14 years; age range 23-67 years) were recruited and signed an informed consent to participate in the study. Participant identifiers were not known to anyone outside the research group. The health status of the participants was assessed through screening, a standard neurological examination performed by a Licensed Independent Practitioner from National Institute of Neurological Disorders and Stroke and a clinical brain MRI. Individuals were excluded if they were deemed to have a medical condition such as neurological disorders, diabetes, uncontrolled hypertension, prior brain surgery or other relevant health issues. All imaging was performed on a 7T Magnetom Terra (Siemens Healthineers, Erlangen, Germany) using a 1Tx-32Rx head array (Nova Medical, Wilmington, USA). No dielectric pads were used.

### 2.2. MRI protocol optimization

The MRI sequence parameters that were developed and implemented are summarized in Table 1. CSF-to-tissue CNR of a high-resolution (0.5 mm isotropic spatial resolution or 0.125 mm^3^ voxel volume) 3D TSE sequence at 7T was optimized using different TEs and echo train lengths (ETLs) while restricting the scan time to ∼10 min. Intra-subject TSE scans were aligned using MATLAB (R2022b, MathWorks, Natick, MA, USA) affine registration tool *imregister.* Signal intensities in CSF ventricles, WM, and deep GM were estimated using four regions of interest (ROIs) of varying sizes drawn in a central slice. The WM and deep GM ROIs were placed in regions excluding PVSs but near the ventricles to minimize contrast variations due to non-uniform B_1_ distribution at 7T. The CNR was calculated as the mean signal difference between CSF and tissue ROIs divided by the standard deviation of pixels within tissue ROIs.

**Table 1:** Protocol sequence parameters for 7T MRI scans conducted in this study. The CSF-to-tissue CNR of 3D TSE was assessed at three varying TEs and image sharpness was evaluated at echo train length (ETL) 108 and 151 using 4.64 ms echo spacing (ES), respectively corresponding to a ETL duration of ∼500 ms and ∼700 ms. Optimal TSE parameters are highlighted in bold. A high-resolution MP2RAGE was acquired to segment brain tissues and cortical areas with a high accuracy. A FLAIR was also performed to detect potential WMH and lacunes that may be confounded with PVSs.

|  | **TSE** | **MPRAGE** | **FLAIR** |
| --- | --- | --- | --- |
| **Orientation** | sagittal | sagittal | sagittal |
| **Matrix size** | 400 x 400 x 320 | 454 x 480 x 320 | 272 x 272 x 224 |
| **Resolution (mm<sup>3</sup>)** | 0.5 x 0.5 x 0.5 | 0.5 x 0.5 x 0.5 | 0.74 x 0.74 x 0.74 |
| **Refocusing FA (°)** | 100 | - | variable |
| **TR (ms)** | 3400/**2430** | 5000 | 9000 |
| **TE (ms)** | 300/**500**/700 | 1.85 | 273 |
| **TI (ms)** | - | 740/2700 | 2300 |
| **BW (pixel/Hz)** | 500 | 360 | 681 |
| **ES (ms)** | 4.64 | 7.3 | 3.9 |
| **ETL** | 151/**108** | 264 | 165 |
| **IPAT (GRAPPA)** | 4 | 3 | 6 |
| **Slice partial Fourier** | - | 6/8 | - |
| **Elliptical scanning** | yes | no | no |
| **TA (min:sec)** | 10:45 | 14:00 | 10:00 |

Numerical simulations based on an Extended Phase Graph algorithm (Hennig et al., 2003) were conducted to assess CSF and tissue signal evolution across varying TEs, refocusing flip angles (FA) and TRs (Fig S1). Additionally, a high-resolution T_1_-weighted MP2RAGE (Marques et al., 2010) was acquired for tissue segmentation and a fluid attenuated inversion recovery (FLAIR) (Hajnal et al., 1992) was performed to detect any WM hyperintensities (WMH) or lacunes.

### 2.3. Processing pipeline and PVS quantification

An outline of the semi-automated PVS processing framework is illustrated in Figure 1. The MP2RAGE and FLAIR images were aligned to the TSE images using a linear affine transformation with *3dAllineate* in AFNI (R. W. Cox, 1996). Alignment was guided using a combination of the local Pearson absolute (lpa) and lpa+ZZ cost functions, which apply a combination of non-linear local weightings to derive matches based on the global set of details (Saad et al., 2009). For computational efficiency of aligning the high-resolution datasets, downsampling in place by a factor of 3 in all dimensions was used in the initial alignment, where a larger phase space of parameters is searched, with subsequent refinement at full resolution. The first inversion MP2RAGE images were aligned to the TSE images, and the same transformation was applied to the second inversion and UNI images, which combine the first and second inversion dataset. Next, the FLAIR image was aligned to the already aligned second inversion image.

**Figure 1:**
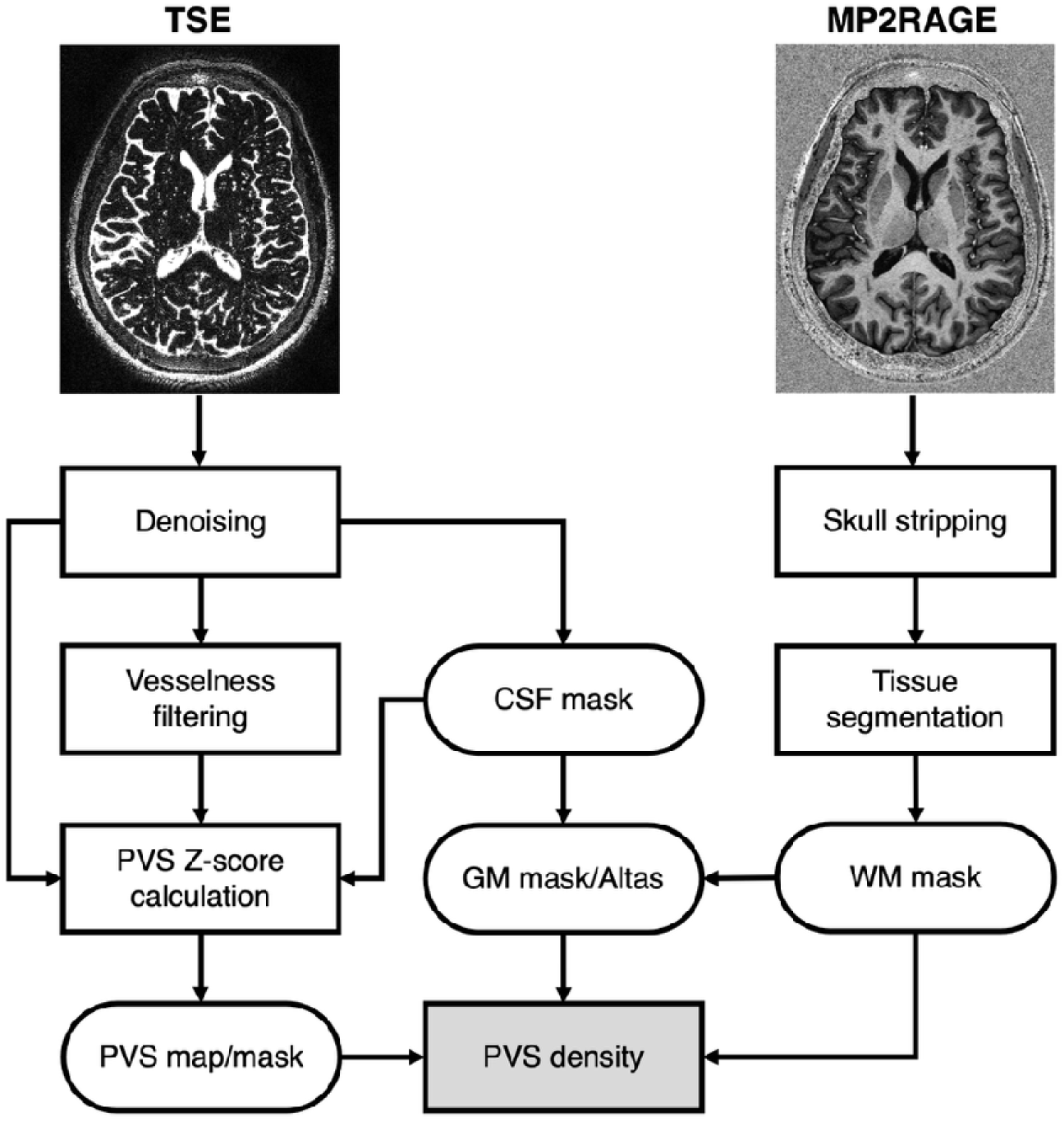
Flow chart of semi-automated PVS processing. PVS extraction consisted of applying a vesselness Frangi filter to the denoised TSE images and calculation of a Z-score map of each vessel-like structures based on their average surrounding noise. In addition, the TSE images were used to extract a bulk CSF mask while skull stripping and WM segmentation was conducted on MP2RAGE high-resolution anatomical images with FSL. GM mask was generated by extracting the region between WM and bulk CSF masks. A cortical atlas was then generated using GM mask. PVS density was calculated by the ratio of PVS voxels detected with a Z-score > 2.58 in tissue mask divided by the total number of voxels in the tissue mask or the cortical area. Note that intracortical PVS structures were excluded from PVS masks, retaining only detected cortical PVS voxels intersecting any PVS clusters connected to WM.

For tissue segmentation, MP2RAGE second inversion datasets were skull stripped using FSL-BET (Brain Extraction Tool) (Jenkinson et al., 2005) to produce a brain mask. The brain-masked T_1_-weighted MP2RAGE UNI images were used to generate a WM mask using FSL-FAST (FMRIB’s Automated Segmentation Tool) (Y. Zhang et al., 2001). A flood-fill operation was performed on the WM mask to remove any imperfections arising from large WM PVSs mistaken as GM tissue during segmentation. The heavily T_2_-weighted TSE images were denoised by applying a block-matching 4D filtering (BM4D) to further improve the CNR and ease CSF/PVS segmentation (Maggioni et al., 2013). A bulk CSF mask was extracted from the denoised TSE images by adaptive thresholding using the MATLAB function *adaptthresh* with a cubic neighborhood of 35 voxels and selection of largest connected cluster with MATLAB function *bwconncomp.* Dilatation of 1 voxel was applied to the CSF mask to exclude potential residual partial volume contamination of bulk CSF signal with tissue. The GM mask was generated by extracting the region between the WM and CSF masks. Additionally, a multi-modal cortical parcellation atlas with 180 areas per hemisphere (Glasser et al., 2016) was aligned to MP2RAGE UNI images and refined using AFNI’s *3dROIMaker* and *3dcalc* to ensure ROI boundaries after mapping were constrained within the subject’s GM mask and that resampling left no gaps among ROIs. Nonlinear alignment from the MNI 2009c asymmetric template was performed using AFNI’s *SSwarper* program, which wraps around *3dQwarp* (R. Cox & Glen, 2013). Again, results of alignment were checked visually for quality, and the estimated transformation was applied to map the reference atlas to each subject’s TSE image space. The separation of the cortex and deep GM was performed by removing the atlas region from the GM mask.

PVS mapping consisted of calculating the Z-scores in each vessel-like structure detected in the denoised TSE images based on their local noise in a surrounding region. The vesselness of each voxel was estimated by 3D Frangi filtering (Frangi et al., 1998) using default parameters (*α*=0.5, *β*=0.5, c=500). The Frangi filter scale was set to a large range of 1 to 11 voxels with a step size of 1 voxel to maximize the inclusion of potentially very small PVSs. To estimate the local noise, a 3D spherical region with 2 mm radius was defined around each potential PVS by applying a morphological dilation in 3D, excluding any overlapping vessel-like structures within this region. The local Z-score was calculated as the signal difference between the potential PVS signal and the mean signal of the surrounding region, divided by the standard deviation of the signal within that surrounding region. The Z-score map was thresholded at 2.58, corresponding to the 99.5^th^ percentile of one-sided Gaussian distribution, to generate the PVS mask. This threshold was empirically determined based on visual inspection and provided a conservative local CNR criterion ensuring that the PVS voxel signal intensity level was clearly above the mean local noise. Subsequently, non-PVS voxels included in the PVS mask, such as residual bulk CSF and potential WMH or lacunes, were manually removed. Isolated intracortical PVS structures were also excluded because they were small which made reliable identification as vessel-like structures difficult. Whole-brain PVS clusters were identified using *bwconncomp*, and WM PVS clusters that intersected any cortical PVS voxel were retained in full and classified as PVS with a leukocortical segment.

The PVS densities in the WM, deep GM, and cortical GM masks were calculated as the percentage of the number of voxels in the PVS masks divided by the total number of voxels in the tissue masks. We then calculated the percentage of WM PVS clusters and WM PVS volume fraction with a leukocortical segment. The region below the top of cerebellum commonly affected by signal reduction due to the use of head arrays at 7T was excluded from the calculation. Scan-rescan data were acquired in 7 subjects within the same session without repositioning in the magnet. The reproducibility of PVS density measurements was assessed by calculating the intraclass coefficients (ICC) using one-way random effects model (1,1) and by Bland-Altman analysis. The PVS density was also quantified in 266 cortical areas (in both hemispheres) of the brain atlas within the brain region selected for analysis.

## 3. Results

### 3.1. TSE parametrization for optimal PVS detection

Figure 2A shows heavily T_2_-weighted TSE images acquired at varying TE values for a representative subject; corresponding simulations are provided in Fig. S1A-C. A reduced and constant refocusing flip angle was found superior to variable flip angle approaches to reduce blurring due to shorter ETL needed to maximize the CNR within our scan time limit of 10 min (Fig. S1D-F). This resulted in a sharper point spread function, effectively enhancing the visualization of small PVSs. CSF-filled PVSs can be identified as hyperintense punctate or tubular structures, depending on their orientation in presented axial images. As predicted by simulations, TSE images acquired at TEs of 500 and 700 ms using a ETL of 151 (echo train duration 700 ms) exhibited strong CSF signal against a markedly suppressed background signal. Measured CSF-to-tissue CNR (∼100:1) was optimal for both WM and GM at TE = 500 ms (Fig 2B). Maximum intensity projection (MIP) of these images shows elongated features that are PVSs (Fig 2C). Use of a protocol with a shorter ETL of 108 (echo train duration 500 ms) maintaining 500 ms TE improved the overall image sharpness, allowing better depiction of small PVSs. CSF-to-tissue CNR decreased to ∼54:1 with shorter ETL of 108 due to the use of a shorter TR required to maintain scan time under 10 min, as expected from simulations. The CSF-to-tissue CNR of denoised TSE images at shorter ETL was ∼180:1, corresponding to a 230% enhancement compared to the original image.

**Figure 2:**
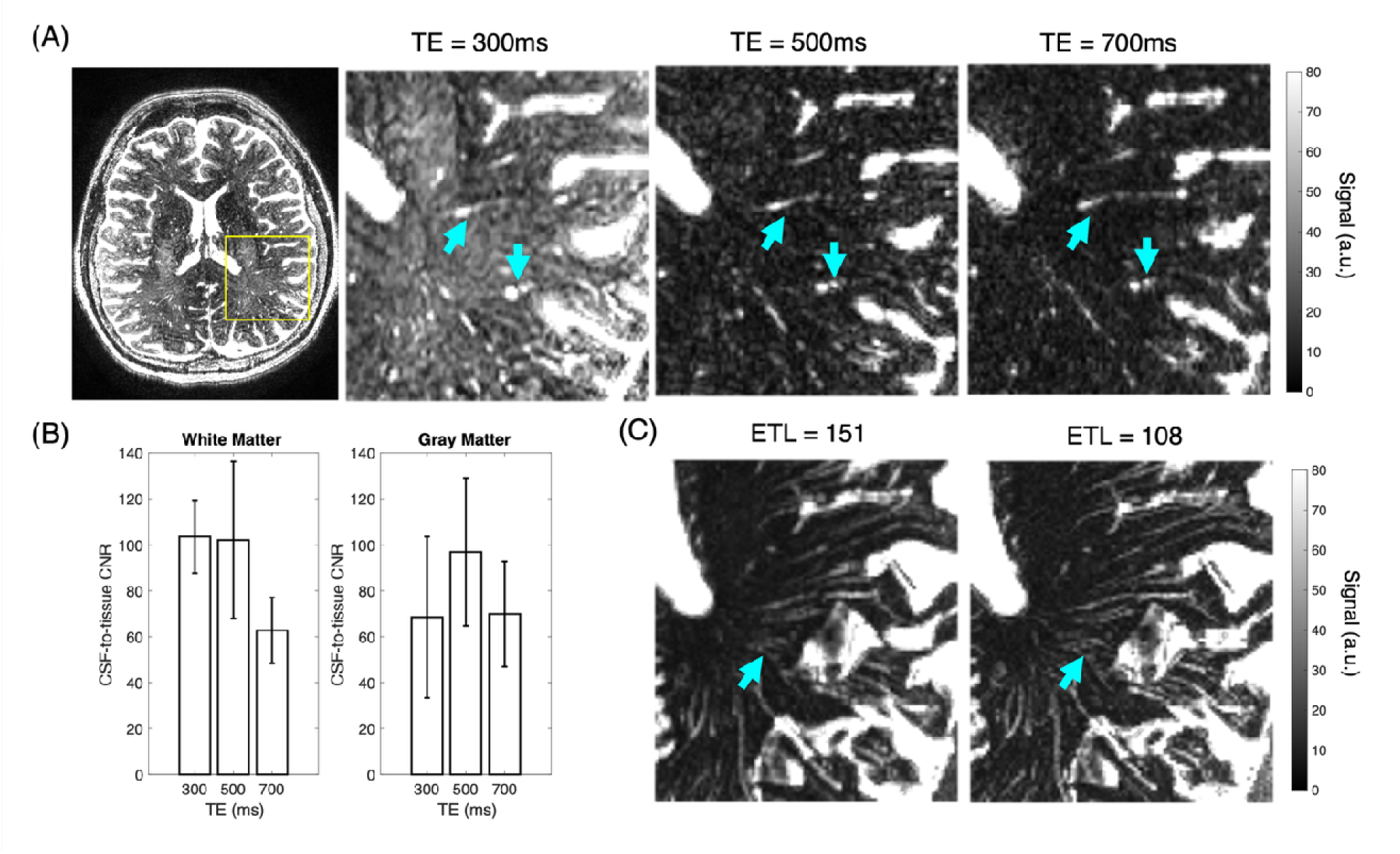
TSE optimization with (A) TSE images acquired at varying effective TEs in a representative subject and (B) corresponding CNR (mean std) determined across four subjects. PVSs can be visualized as bright punctate and linear structures as pointed by the cyan arrows. Note that the contrast is optimal at TE = 500 ms (TR = 3.4 s and ETL = 151) for both the WM and GM. (C) Comparison of 5.5 mm MIP of TSE images (TE = 500 ms) acquired with a ETL of 151 (TR = 3.4 s) and 108 (TR = 2.4 s). PVSs can be identified as linear structures with high signal intensity. Use of a reduced ETL improved the visualization of small PVSs as pointed by the cyan arrows.

### 3.2. Whole-brain semi-automated PVS quantification

PVSs were segmented with the data processing pipeline described in Fig. 1 using TSE images acquired with the optimal parameters listed in Table 1. TSE image (Fig 3A) and its associated thresholded PVS Z-score map (Fig. 3B) identify signal intensities exceeding 2.58 standard deviation above local background noise. This enabled detection of PVS signal with a strong local CNR corresponding to the 99.5 quantile of a Gaussian distribution. The Z-score threshold reflects a conservative local contrast criterion and not a statistical confidence interval.

**Figure 3:**
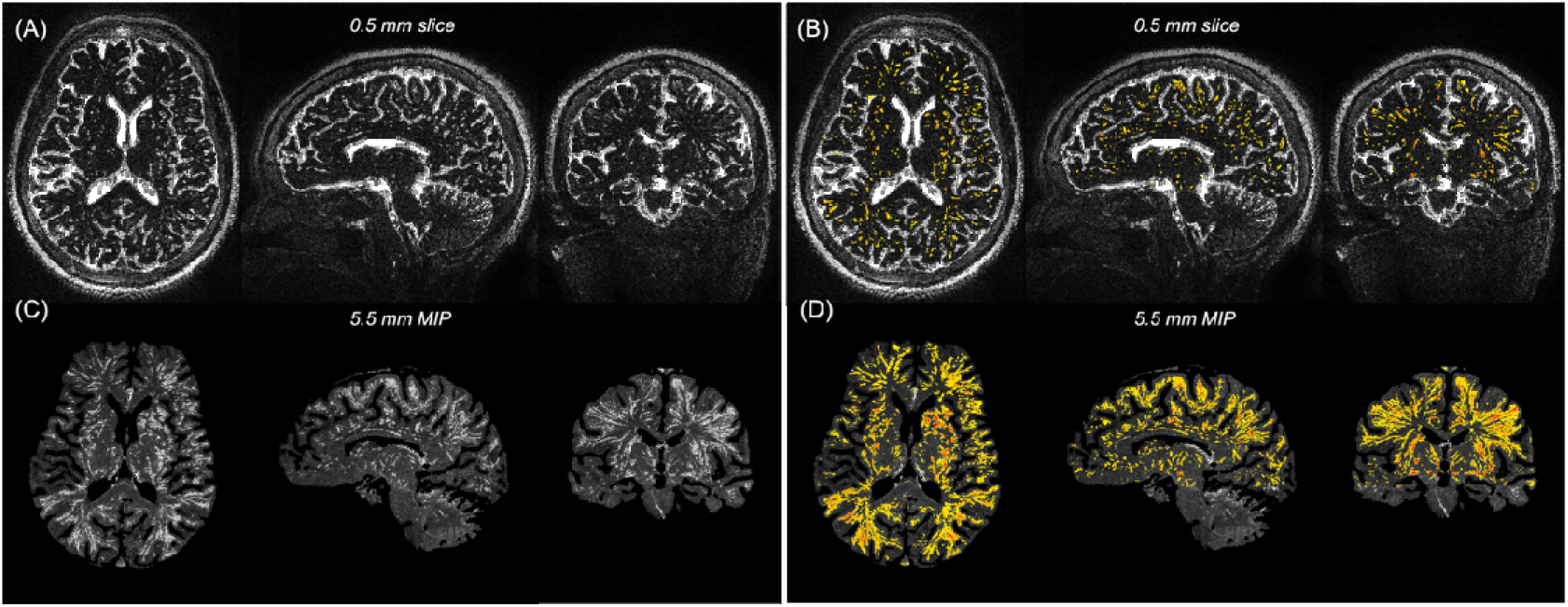
Optimal TSE acquisition with resulting PVS Z-score segmentation. (A) displays the TSE image acquired with optimal parameters listed in Table 1, along with (B) overlay of the Z-score maps (in color Z>2.58) generated with the pipeline presented in Figure 1. As expected, many PVSs appearing as discrete spots in single slice are revealed to be elongated vessel-like structures in (C,D) corresponding maximum intensity projection (MIP) across 11 slices. Note that bulk CSF was removed from MIPs to improve visualization of the PVSs. Moreover, the PVS segmentation was performed excluding the region below the top of cerebellum commonly affected by signal reduction due to the use of head arrays at 7T.

Denoising effectively reduced background tissue noise while preserving CSF and PVS sharp appearance. The noise-only background limited the risk of structure smoothing. An example of the relationship between the TSE signal, before and after denoising, with corresponding PVS Z-score is shown in Figure S2.

PVSs exhibited Z-score values as high as 114±63 across subjects with a mean Z-score of 6±1. Z-scores close to CSF-to-tissue CNR measured in ventricles were likely associated with large PVSs while lower Z-scores were attributed to partial volume effects of small PVSs with surrounding tissues. As expected, most of extracted PVSs appearing as discrete structures were revealed to be elongated vessel-like structures, when viewed in the corresponding MIPs (Fig 3C, D). The overlay of the magnitude gradient of the CSF image is shown on aligned MP2RAGE UNI image for a representative subject in Figure 4A. The edge of the bulk CSF perfectly aligns with the border between CSF and gray matter in the MP2RAGE image, indicating excellent registration of the anatomical image on the TSE image (additional examples in Fig S3). This allowed precise localization of each PVS within anatomical regions, as illustrated by the PVS Z-score map overlaid on the MP2RAGE anatomical image in Figure 4B.

**Figure 4:**
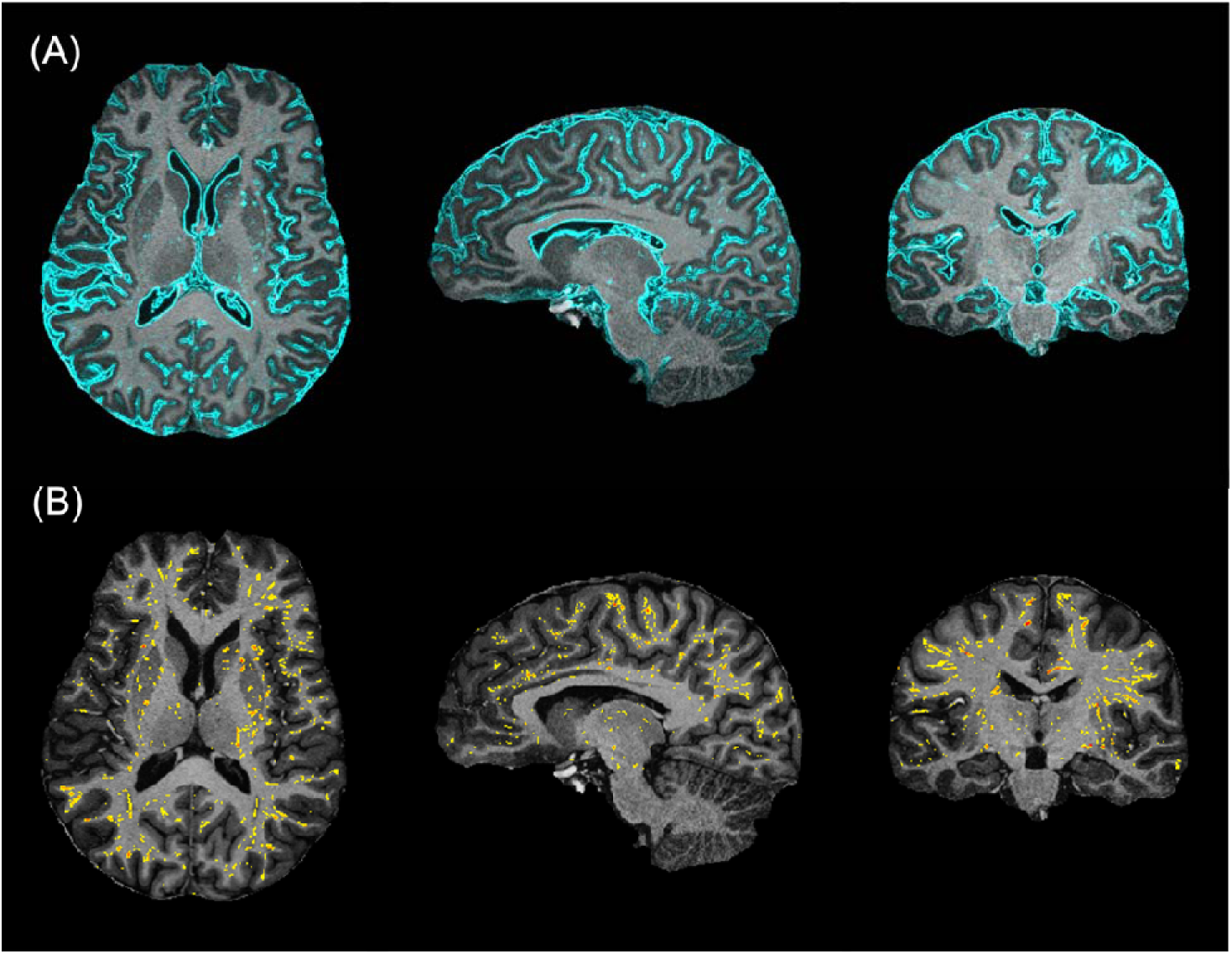
Example of TSE and MP2RAGE alignment with PVS localization in anatomical regions. (A) Overlay of the edges of the TSE image (in cyan) precisely aligns with the interface between the dark CSF border and gray matter in the registered MP2RAGE UNI image, indicating excellent anatomical alignment. (B) PVS Z-score map (in color Z > 2.58) overlaid on the MP2RAGE UNI image, highlighting extracted PVSs position in anatomical regions.

### 3.3. PVS characterization at the gray-white matter interface

Using the acquisition and data processing protocol described above, approximately 20 3% of WM PVS clusters across subjects were found to have a leukocortical segment. The portion of WM PVSs with a leukocortical segment accounted for 74 5% of the total PVS volume. This indicates that WM PVSs connected to the gray-white matter interface accounted for majority of the PVS volume, while a substantial fraction of WM PVSs corresponded to small isolated PVS structures. Figure 5 presents representative examples of PVSs at the gray-white matter interface, likely associated with medullary arteries, across different individuals. Figure 5A and 5B show examples of individual PVSs with a leukocortical segment, while Figure 5C illustrates a case of PVS appearing as a cluster with multiple leukocortical segments within a common cortical area.

**Figure 5:**
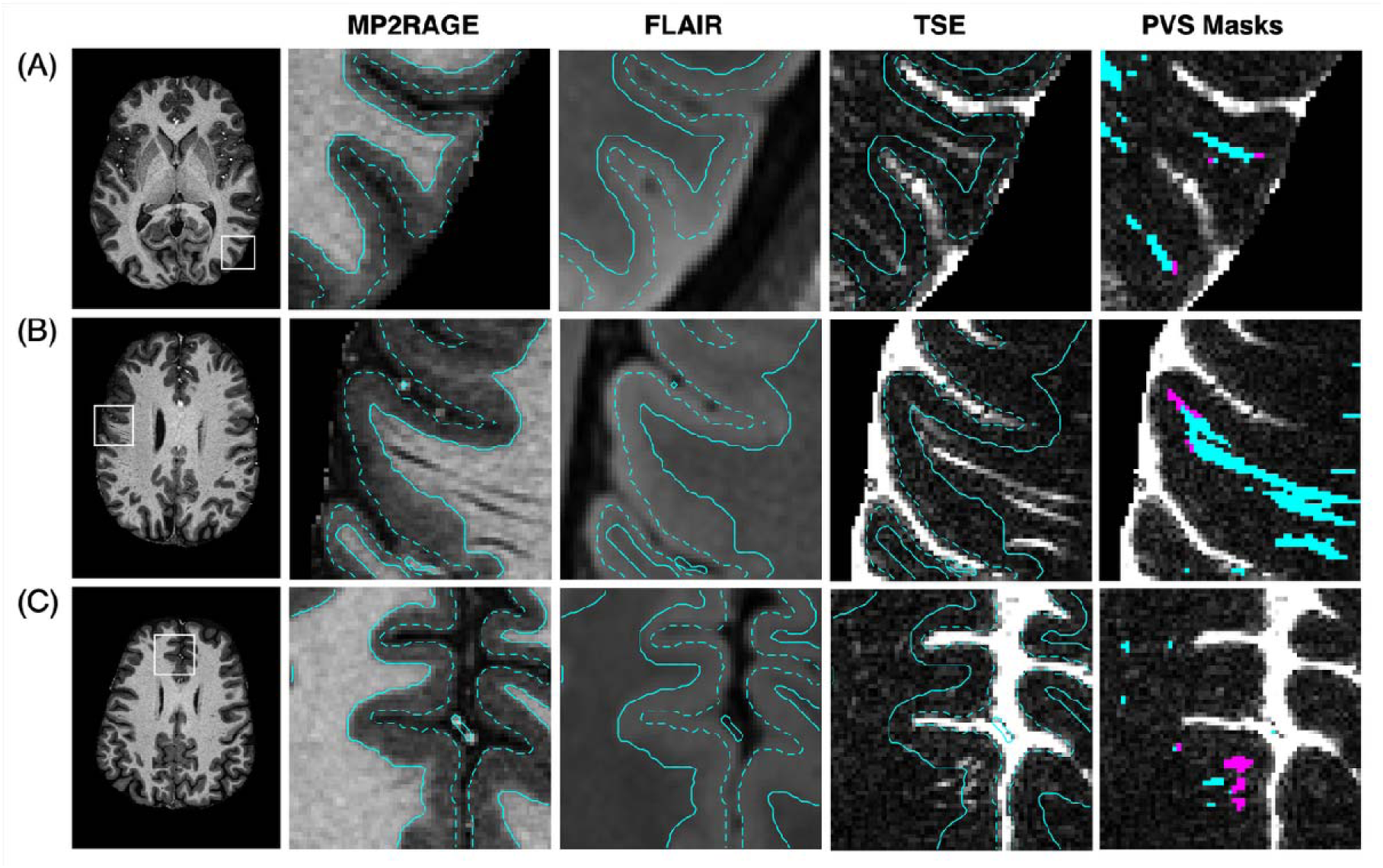
Representative examples of PVSs with a leukocortical segment in the gray-white matter interface. The first column shows anatomical locations on M2RAGE images across different subjects, with corresponding zoomed regions in each row for the UNI MP2RAGE, FLAIR and TSE images. In each scan, the segmented cortical boundary is overlaid as a solid cyan line while the dotted line delineates outer cortical GM surface at the GM-CSF interface. The 5^th^ column shows the PVS masks overlaid on TSE images with WM PVS segments highlighted in cyan and their leukocortical segments in magenta. PVSs with a leukocortical segment were frequently observed in the gray-white matter interface either as individual structure (A,B) or clusters (C). These PVSs were likely associated to medullary arteries originating from small penetrating arterioles in the adjacent cortical areas. Large PVSs at the boundary may have affected the definition of the cortical boundary on MP2RAGES images. Unfortunately, FLAIR was not sufficient to identify the true location of the cortical boundary. Note that the outer surface of the mask did not coincide perfectly with the outer edge of the FLAIR images because of its lower spatial resolution (0.74 mm isotropic resolution) compared to MP2RAGE and TSE (0.5 mm isotropic resolution).

Leukocortical PVS segments get increasingly smaller in the cortex likely due to compression around cortical penetrating vessels, resulting in a Z-score below threshold. The potential connection of PVSs with the subarachnoid space was difficult to assess, notably due to partial volume effects induced by bulk CSF within GM regions. Although the PVS sections in WM can be identified as hypointense on MP2RAGE images (Fig 5, column 2), the connecting leukocortical PVS segments could only be reliably detected with processing of the TSE images (Fig 5 column 5). Most PVSs were not visible on FLAIR images but FLAIR confirmed that PVSs were not confounded by WMH or lacunes. GM mask outer surface did not coincide perfectly with the FLAIR outer edge due to its lower spatial resolution.

Figure 6 presents additional examples of PVSs likely associated with large insular arteries. As expected, the leukocortical segments of these PVSs exhibited high Z-score values (69±16), suggesting the presence of large PVS compartments. The morphology of these PVSs often appeared as dense groups or clusters in both hemispheres, consistent with the anatomy of perforating arteries. A slight denoising-induced smoothing resulted in a patch-like appearance in PVS masks. FLAIR contrast was further affected by B_1_ inhomogeneities in this inferior region but confirm the absence of confounding effects.

**Figure 6:**
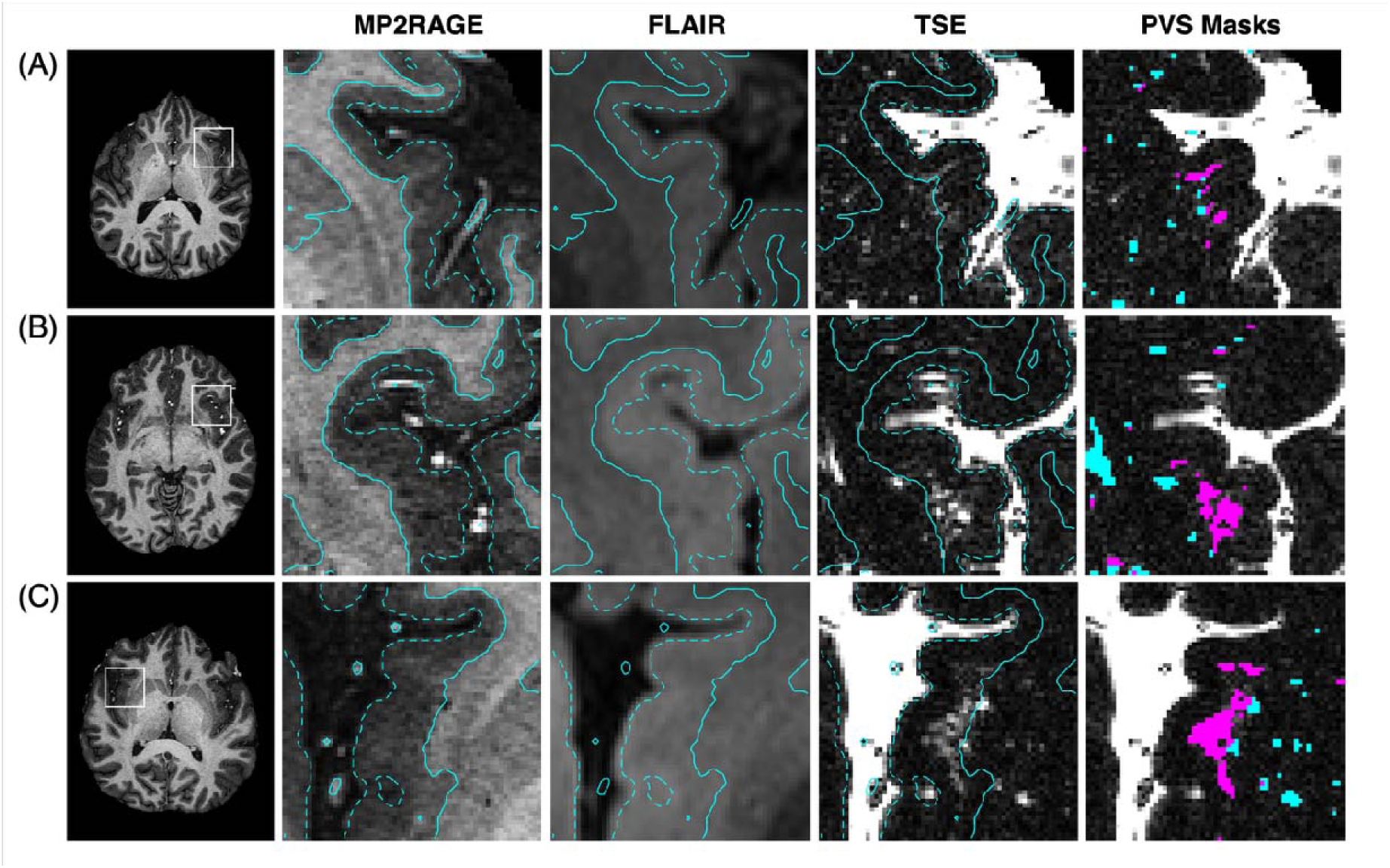
Additional examples of PVSs exhibiting a leukocortical segment in the gray-white matter interface. The first column shows anatomical locations on axial MP2RAGE images acros subjects, with corresponding zoomed regions of the insula displayed in each row. Segmented cortical boundary is overlaid as a solid cyan line while the dotted line delineates outer cortical GM surface at the GM-CSF interface. In this region, PVSs were frequently observed as clusters, likely associated with groups of perforating arteries branching from the MCA. In the fifth column, PVS masks overlaid on TSE images highlight large leukocortical PVS segments in the insula (magenta) associated to WM PVSs (cyan). Note that PVS clusters were systematically found in both hemispheres. Large PVS clusters may further alter T_1_-contrast in MP2RAGE images, affecting cortical boundary delineation. FLAIR contrast was further degraded due to B_1_ inhomogeneities in this inferior region.

In some cases, PVS-induced signal reduction on MP2RAGE images may have locally impacted the definition of the cortical boundary in tissue segmentation, especially for large PVSs and dense PVS clusters (Fig S4). Line-scan analysis across contrasts supported that leukocortical PVS segments presented in Figure 5C have a portion within cortex as signal was observed to spread deeper within the cortical region, towards the CSF-GM interface (Fig S4A). Subsets of leukocortical PVS segments in Figure 6 may have been overestimated due to potential errors in tissue masks. As shown in line-scan analysis of Figure S4C, the precise location of the boundary in this region was hard to assess in both MP2RAGE and FLAIR images, yet PVSs with a lower signal intensity in the TSE likely did not affect WM MP2RAGE signal and appear to be located within the cortex.

### 3.4. PVS density in WM, deep GM, and cortex

The average PVS density was found to be 4.3±1.5% in WM, 2.6±1.7% in the deep GM and 0.74±0.4% in the cortex (Fig S5). The PVS density in the cortex represents only the segments in the gray-white matter interface. The scan-rescan reliability showed good intrasubject reproducibility of repeated measurements with an ICC of 0.93, 0.98, and 0.63 respectively for WM, deep GM, and cortex. The 95% limit of agreement in Bland-Altman analysis was slightly exceeded in two subjects in the WM, likely due to motion artifacts in rescan TSE acquisitions. PVS density in WM and deep GM varied with age (Fig S5A), whereas it appeared to be more stable in the cortex.

Figure 7 shows the 20 cortical areas with the highest PVS density measured across the 17 subjects. Among 266 specific cortical areas analyzed above the cerebellum, top PVS densities (∼2.7-6.2%) were found related to the anterior agranular insula complex (AAIC) and the posterior insular areas (PoI1, PoI2) supplied by the branches of the MCA. The primary auditory cortex (A1) and the lateral belt complex (LBelt) were found to have lowest PVS densities (<0.04 %). Table S1 summarizes all regional PVS densities of the 266 cortical areas averaged across all the participants.

**Figure 7:**
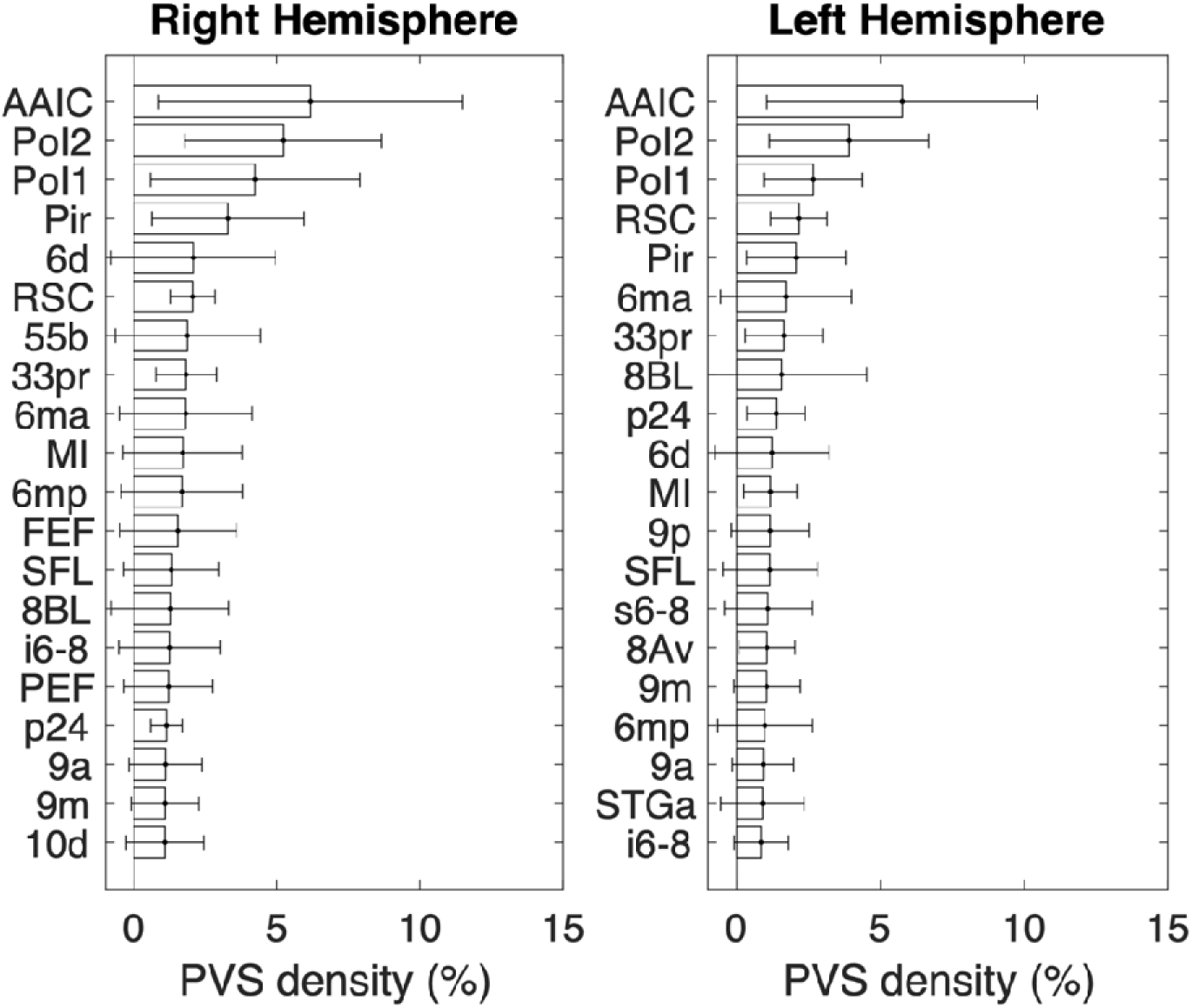
Twenty highest cortical PVS density in atlas regions for both hemispheres. Highest PVS density (∼3-6%) was found in anterior agranular insula complex (AAIC) and posterior insular areas (PoI1, PoI2) while lowest (< 0.04%) were found in primary auditory cortex (A1) and the lateral belt complex (LBelt).

## 4. Discussion

This study introduces an optimized heavily T_2_-weighted 3D-TSE sequence at 7T that achieves high CSF-to-tissue CNR while minimizing background tissue signal, enabling high-resolution whole-brain imaging of PVSs in the gray-white matter interface. Leukocortical segments associated with a majority of WM PVS volumes were readily detected, as the TSE contrast was tailored to maximize CSF-to-GM CNR. Using the semi-automated PVS processing framework developed here, PVS density was quantified across individual cortical areas throughout the brain, allowing assessment of leukocortical PVS burden in the gray-white matter interface.

Brain tissue suppression with TSE was excellent at TE=500 ms when using a constant refocusing flip angle of 100° and a TR of 2.4 s. Employing a short ETL and reduced flip angle resulted in lowering the echo signal variation across the ETL, leading to minimal blurring as well as lower SAR. Reduced and constant refocusing flip angle was found to increase image sharpness compared to variable flip angle strategies, such as sampling perfection with application-optimized contrasts using different flip-angle evolutions (SPACE) (Mugler et al., 2000), which required extended ETL to achieve similar CNR. A very high CSF-to-tissue CNR of 180:1 at 0.5 mm isotropic spatial resolution was achieved by combining optimal TSE parameters with denoising, facilitating the PVS quantification in the GM cortical regions. The semi-automated processing method developed in this study relies on the vesselness filtering and calculation of PVS local Z-score. Use of a local Z-score accounts for spatial variation of noise well known to be prominent at 7T (Maximov et al., 2012). A Z-score threshold of 2.58 enabled extraction of PVSs with conservative local CNR criterion providing confidence that each PVS signal intensity was clearly above the mean local noise.

The majority of the leukocortical PVS segments detected were likely associated to medullary arteries as described previously (Kwee & Kwee, 2007). These PVSs were sometimes organized in dense clusters at the gray-white matter interface across various anatomical locations (Fig S4). Groups of large PVSs with leukocortical PVS segments were consistently found within the insula, associated to M2 branches of the MCA, which involve larger arteries that abuts and indents the cortex (McArdle et al., 2020; Varnavas & Grand, 1999). Alignment of TSE images with high-resolution angiography will provide a better understanding of their association with the underlying cerebral vasculature. Intracortical PVSs were occasionally detected by Frangi filtering, however, they lost their vessel-like appearance after Z-score thresholding due to their small size. Therefore, it was difficult to identify these PVSs reliably. Future improvements in MRI sensitivity will be necessary to assess intracortical structures. PVS detection may be enhanced by restoring the longitudinal magnetization between TRs, implementing zero-filling approaches and minimizing background tissue noise further along using advanced denoising methods. These strategies and application of prospective motion correction techniques to reduce image artifacts will be investigated in future studies.

The average WM PVS density (∼4.3%) was found to be within the range reported in previous studies (Barisano et al., 2021; Cai et al., 2015). The cortical PVS density of segments connected to WM PVSs was about ∼0.7% which was, as expected, much lower compared to WM PVS density. However, the actual cortical PVS density may be higher since possible intracortical PVSs were not included in the quantification. The highest PVS density was found in the insula (AAIC, PoI1 and PoI2) while lowest density was found in the auditory cortex (LBelt and A1). In agreement with previous studies (Chen et al., 2011; Zhu et al., 2011), PVS density within WM and deep GM appeared to increase with age. However, the PVS density in the cortex was found to be stable across individuals. Since the PVS density has been found to depend on multiple factors even in the healthy population (Barisano et al., 2021), a larger cohort will be required to robustly assess age-related PVS volume changes.

The brain MRI alignment and segmentation pipeline allowed for the extraction of bulk CSF, WM, and GM as well as the precise definition of 266 cortical areas for the quantification of the PVS densities. Large PVSs or dense PVS clusters near the cortical boundary could affect local boundary delineation due to signal reduction in WM on MP2RAGE images (Fig S4). Therefore, some extracted leukocortical PVS segments may include a greater contribution from WM. This effect may lead to an overestimation of the PVS density in some cortical regions. Although better definition of the boundary would help precisely isolate the cortical segments of these leukocortical PVSs, many PVSs in the gray-white matter interface were observed deeper within cortex, supporting the presence of a distinct cortical segment. A FLAIR protocol with improved gray-white matter contrast may provide additional insight into the definition of the boundary between the tissues. Moreover, due to reduced MP2RAGE contrast at 7T, part of the thalamus was not properly segmented and was included into WM mask, which may have affected deep GM quantification. MRI contrast may be improved with parallel transmission RF pulses at 7T (Gras et al., 2017). This will improve tissue and PVS segmentation in inferior brain regions while minimizing TSE contrast variability across the brain, which should enable quantitative volumetric PVS mapping.

The amount of CSF in any PVS voxel can be estimated by using the relationship that the TSE signal intensity of a 100 % CSF voxel corresponds to a voxel volume of 125 nL (0.125 mm^3^). The mean CSF signal measured within fully CSF-filled voxels in the ventricles was 330±51 across subjects, while the mean background tissue signal was 8.3±1.8. For chosen detection threshold (Z=2.58), the corresponding signal intensity was ∼13 (8.3+2.58*1.8). This threshold resulted in CSF detection limit of approximatively 5 nL (125 nL*13/330). Potential effects such as T_2_ may also influence PVS signal and will have to be accounted for accurate PVS volumetry.

In conclusion, this study optimized a high-resolution (0.125 mm^3^ voxels) T_2_-weighted 3D-TSE sequence at 7T providing a CSF-to-tissue CNR of ∼180:1 with minimal tissue contribution. In addition, this work developed a processing pipeline enabling robust PVS segmentation and quantification in the whole brain. PVSs with a leukocortical segment were consistently identified at the gray-white matter interface in healthy individuals and their density was analyzed across multiple cortical regions. Changes in morphology or density of leukocortical PVS segments may reflect properties of the associated cortical areas and could help characterize various neurological conditions.

## Data Availability

Data collected are available online on https://figshare.com/s/6d97007e79a12b7bc47d. The scripts used in the manuscript are available upon reasonable request.

## Abbreviations

PVS: perivascular space
CSF: cerebrospinal fluid
TSE: turbo spin echo
ETL: echo train length
ES: echo spacing
FA: flip angle
BW: bandwidth
TI: inversion time
TA: acquisition time
TR: repetition time
TE: echo time
WM: white matter
GM: gray matter
CNR: contrast-to-noise ratio
SAR: specific absorption rate

## Data and Code availability

The dataset, including high-resolution 7T MRI scans (T_2_-weighted 3D TSE, MP2RAGE and FLAIR) from 17 healthy participants, as well as retest data from 7 participants, and associated PVS masks, is available at https://figshare.com/s/6d97007e79a12b7bc47d. Computer codes for image alignment and segmentation are available at https://afni.nimh.nih.gov. Frangi filter implementation used for PVS extraction is available on MATLAB File Exchange (https://www.mathworks.com/matlabcentral/fileexchange/24409-hessian-based-frangi-vesselness-filter). Other scripts used in the manuscript are available upon request.

## Author Contributions

GS and ZHD have equal first authorship. ZHD conceived of the presented idea and developed the theory, as well as leading subject recruitment. GS performed technical design, optimizing MRI sequences and performed majority of the data processing. GS conducted the experiments and data acquisitions with ZHD. GS and ZHD analyzed and interpreted the data together. GS wrote the manuscript, with contributions from ZHD. PAT handled MRI image alignments and provided input on technical aspects. SLT provided technical insight on MRI methods, contributed to data analysis and writing of manuscript. APK provided insights on study design and data processing, supervised the project and contributed to the manuscript.

## Supplementary Material

**Figure S1:**
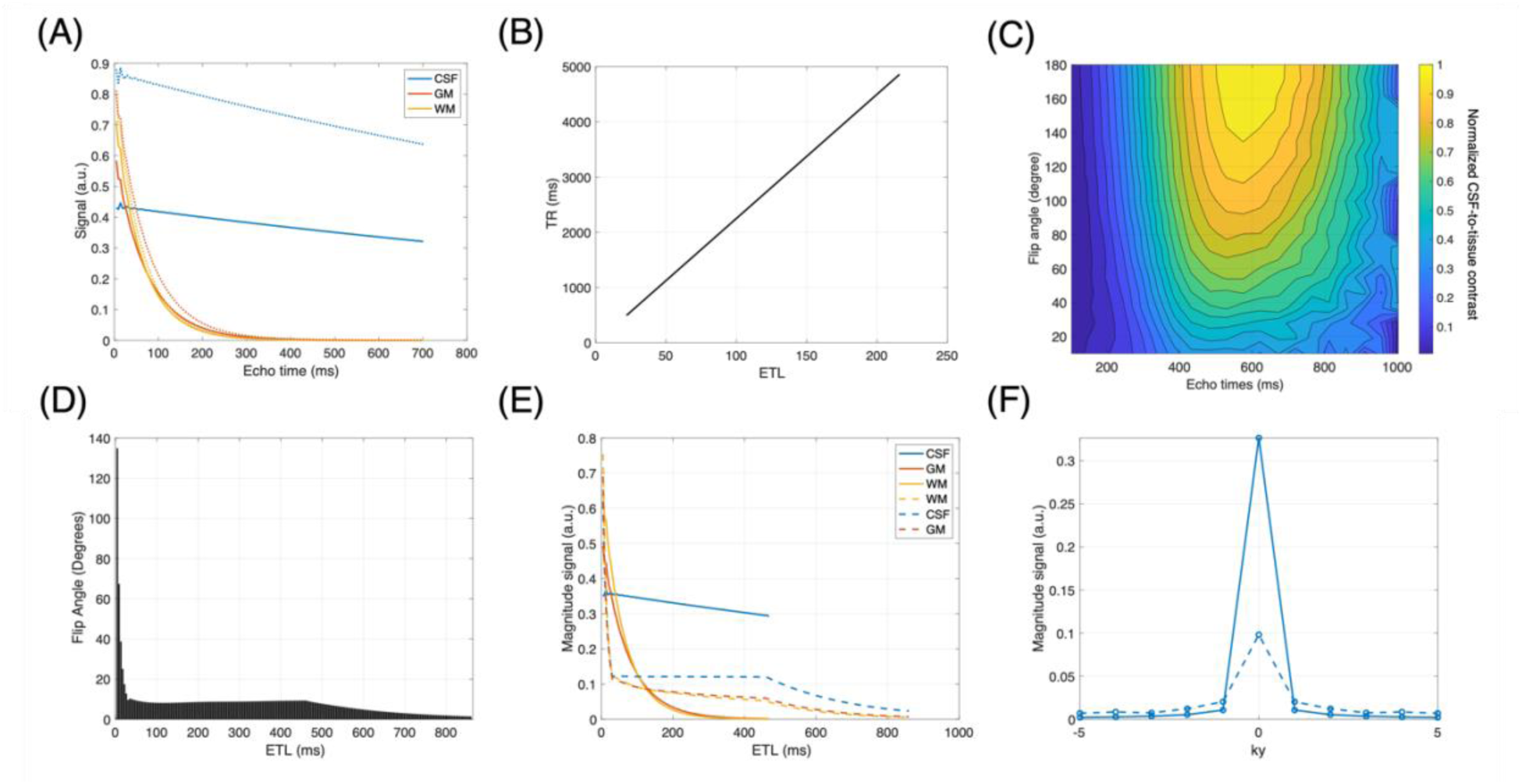
T_2_-weighted MRI sequence simulations using extended phase graphs algorithm. (A) Signal evolution of CSF and tissue for a constant refocusing flip angle (FA) with an echo train length (ETL) of 151 (∼700 ms), for a TR of 3400 ms (solid lines) and TR infinite (dotted line). Achievable FA and echo spacing (ESP) were respectively restricted to 100° and 4.24 ms due to SAR limits. (B) TR as a function to ETL for a total acquisition time (TA) of 10 min, defined as TR(s) = *TA*. *ETL*⁄(*N*_*y*_. *N*_*z*_)/*IPAT*. Elliptical scanning reduced scan duration by ∼ 120 sec. (C) Contour plot of normalized CSF-to-GM contrast as a function of refocusing FA and TE. (D) Variable refocusing FA (VFA) along ETL was extracted from Siemens SPACE sequence using CSF T_1_ = 4300 ms and T_2_ = 2000 ms, with desired sequence requirements. In this case, the ETL and ESP were respectively adapted to 198 and 4.34 ms to achieve a TE of 500 ms, while TR was set to 4.5 s to limit total TA to ∼10 min. (E) Signal evolution of CSF and tissue for constant refocusing FA (solid line) and VFA (dotted line). The use of a constant refocusing FA enables shorter ETL to suppress background tissue signal more effectively compared to VFA. At 500 ms TE, the TSE led to an increase in CSF signal, contrast, and sharpness, as illustrated in (C) by resulting point spread function, where TSE is represented by the solid line and SPACE by the dotted line.

**Figure S2:**
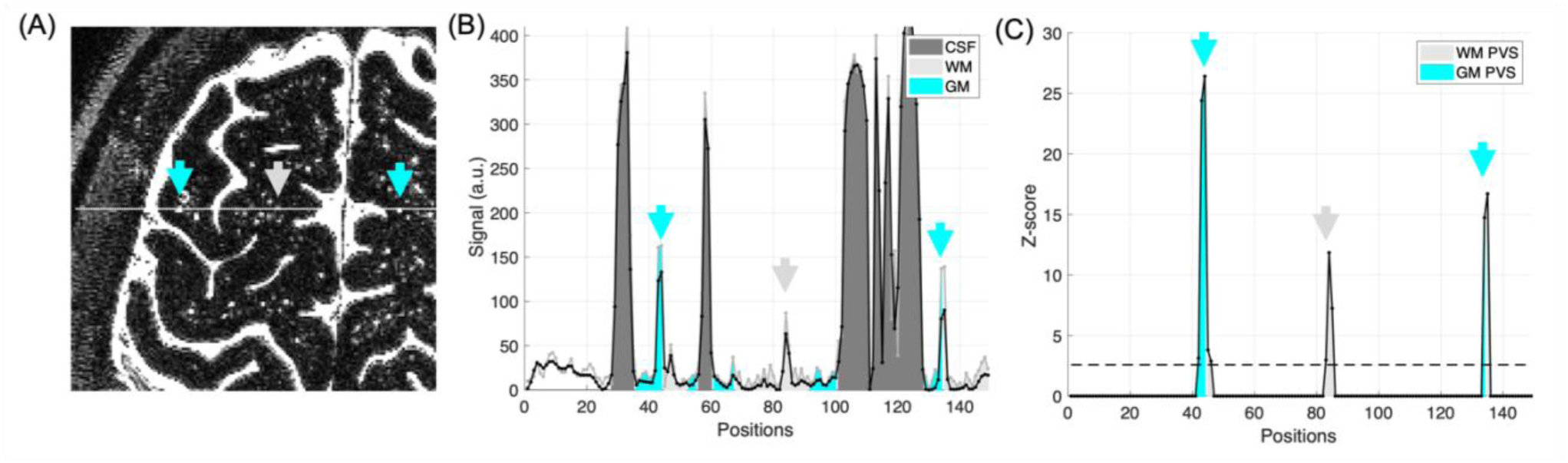
PVS signal intensity profile and corresponding Z-score. (A) A line profile (white line), drawn on TSE image, intersects two leukocortical PVS segments (cyan arrows) and one WM PVS (light gray arrow). (B) TSE signal intensity along this line (black line) shows peaks corresponding to bulk CSF and PVSs. Denoising effectively reduced tissue-related noise while largely preserving the CSF signal (light gray line). Despite a mild smoothing effect of PVS signal, denoising enabled CSF-to-tissue CNR to improve by ∼230%. (C) Corresponding Z-score profile highlights detected PVSs with a Z>2.58. Note that signal peaks not visible in the Z-score plot were either not identified by the Frangi filter or were below the Z-score detection threshold (dotted line).

**Figure S3:**
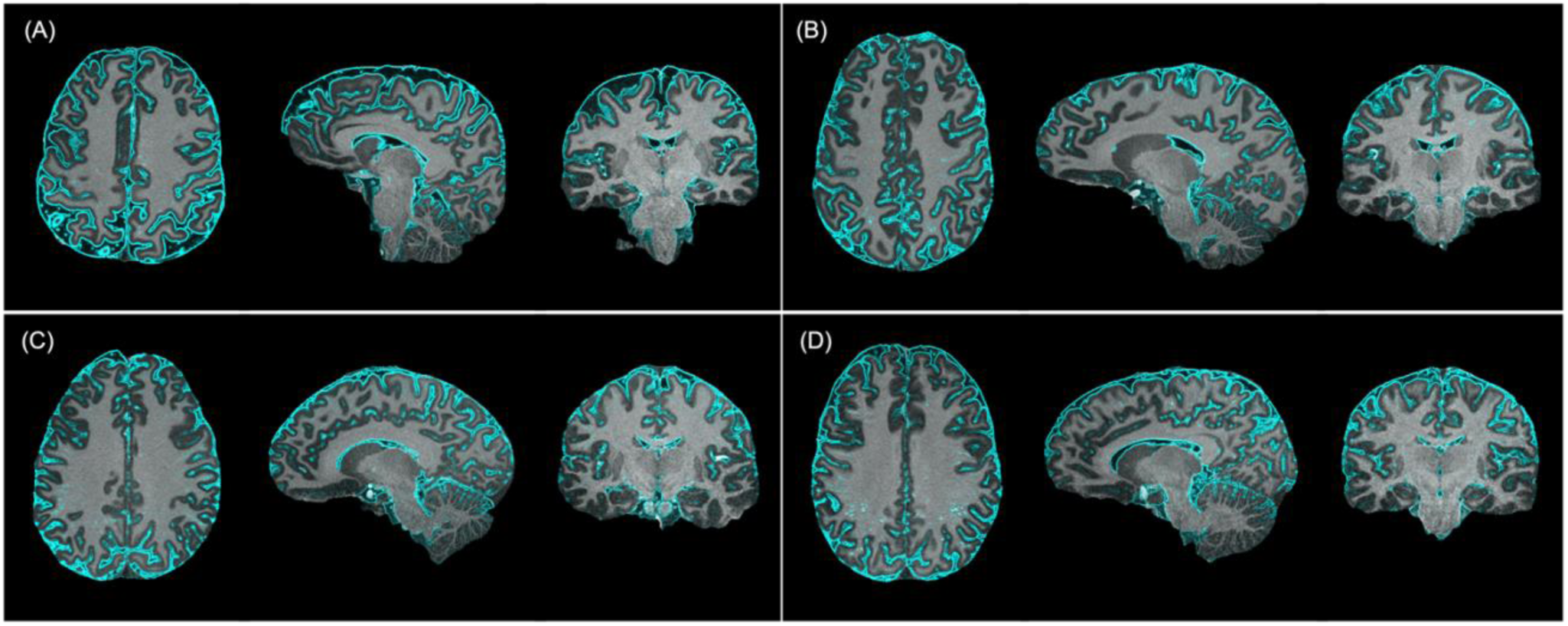
Representative examples showing the alignment of MP2RAGE images to TSE images in 4 additional subjects. Rigid-body registration was performed using AFNI’s *3dAllineate*. The cyan contour overlaid on the MP2RAGE images highlights CSF magnitude gradient from the TSE images and aligns precisely with the interface between the dark CSF border and gray matter in the MP2RAGE scans. This demonstrates the high accuracy of the image registration confirming a strong alignment between PVSs and anatomical regions.

**Figure S4:**
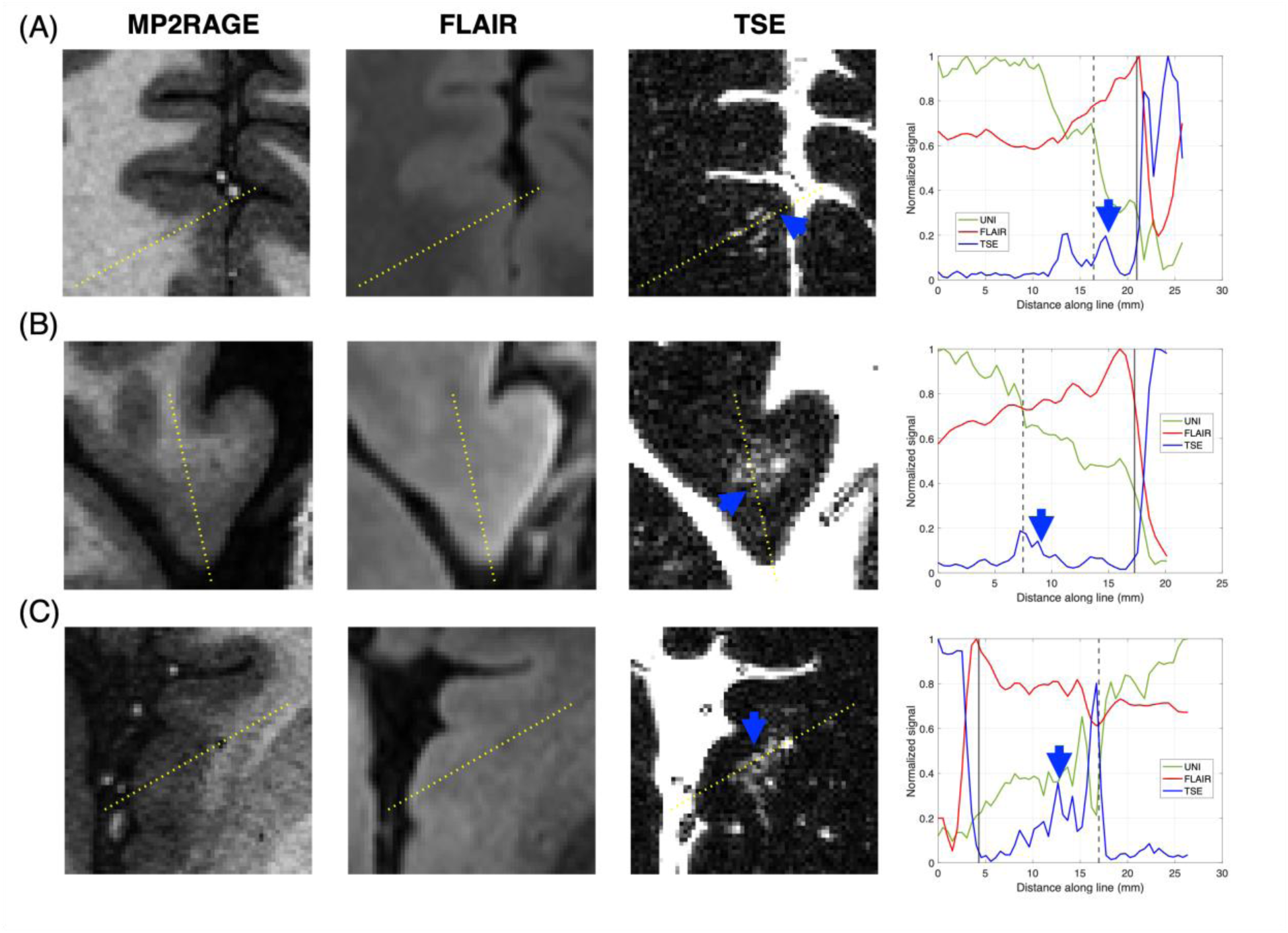
Effect of PVS clusters on cortical boundary delineation in different anatomical locations. MP2RAGE UNI images (first column) show localized blurring of the cortical boundary in areas containing PVSs, visible on TSE images (third column), likely associated to medullary arteries (A-B) and insular arteries (C). PVS structures could not be identified clearly on FLAIR images (second column) and assessment of tissues in these regions was difficult due lower resolution and reduced contrast at 7T. According to MP2RAGE segmentation, these PVS clusters had a subset within cortex (blue arrows). In graphs, normalized line scans along yellow dotted lines in each image show corresponding signal intensity evolution in these areas (from left to right). The black dotted vertical lines represent the segmented cortical boundary while the solid vertical lines correspond to the outer surface of the GM mask at the CSF-GM interface. MP2RAGE signal decreases from the cortical boundary towards the CSF-GM interface, while FLAIR signal tends to increase almost linearly across this region. MP2RAGE signal effect is consistent with presence of large PVSs while FLAIR signal appears insensitive to these PVSs. Although large PVSs may have artificially displaced the boundary into WM, small subsets of these PVSs segments deeper into cortex have limited impact on the MP2RAGE signal. Boundary PVSs exhibited higher signal, with gradual decrease toward the cortex, suggesting a likely cortical portion.

**Figure S5:**
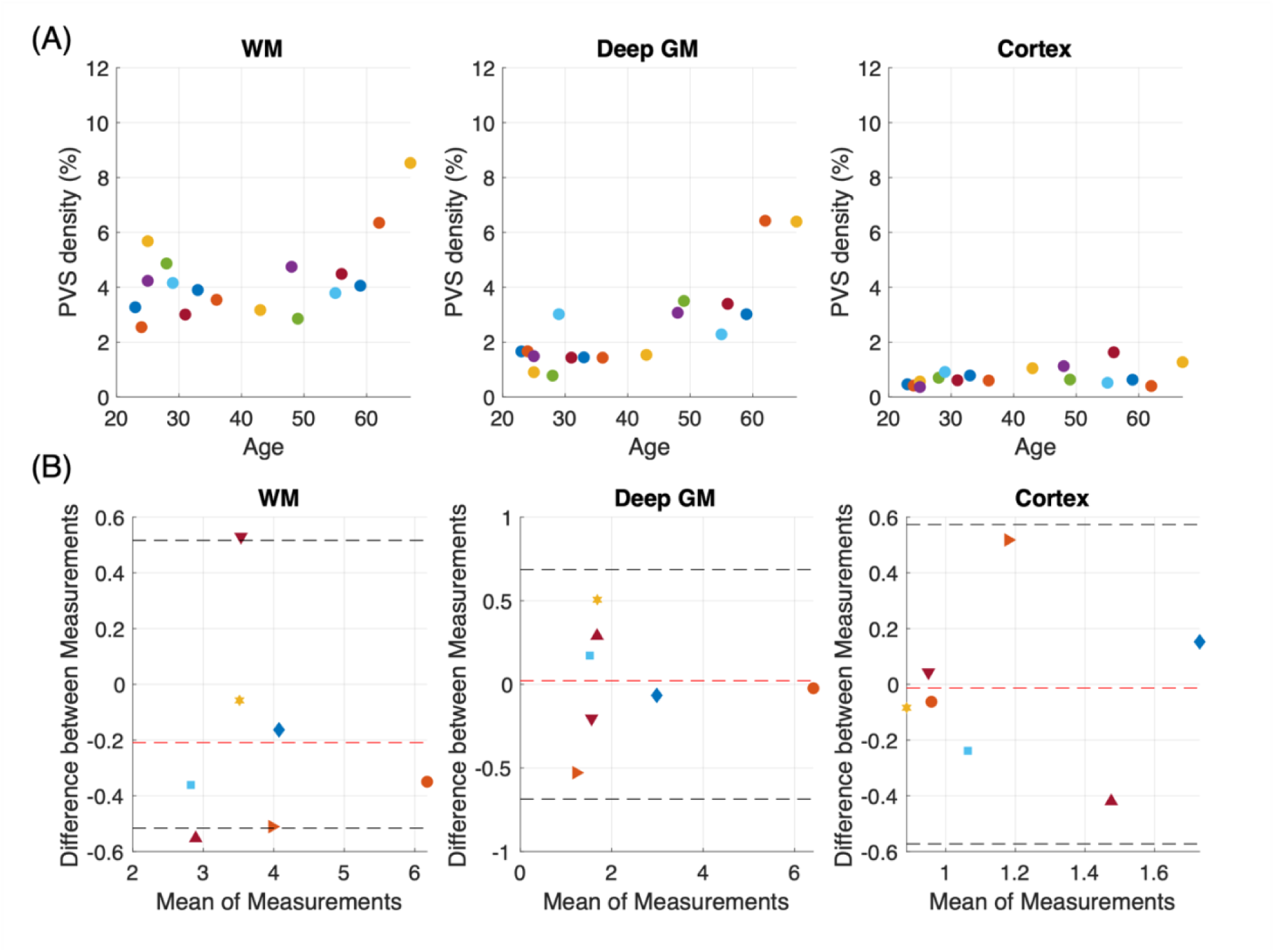
PVS quantification with (A) average density extracted in the 17 subjects within WM, deep GM and cortex with (B) Bland-Altman analysis showing the reproducibility of PVS density measurements in these regions for 7 subjects. The black dotted lines represent 95% interval of confidence while the red dotted line shows the mean of the difference between scan and rescan measurements for each subject. Note that the differences in density found between the scans and rescans slightly exceeded 95% confidence interval in the WM for two subjects. This difference was due to motion artefacts within the second scan.

**Table S1:**
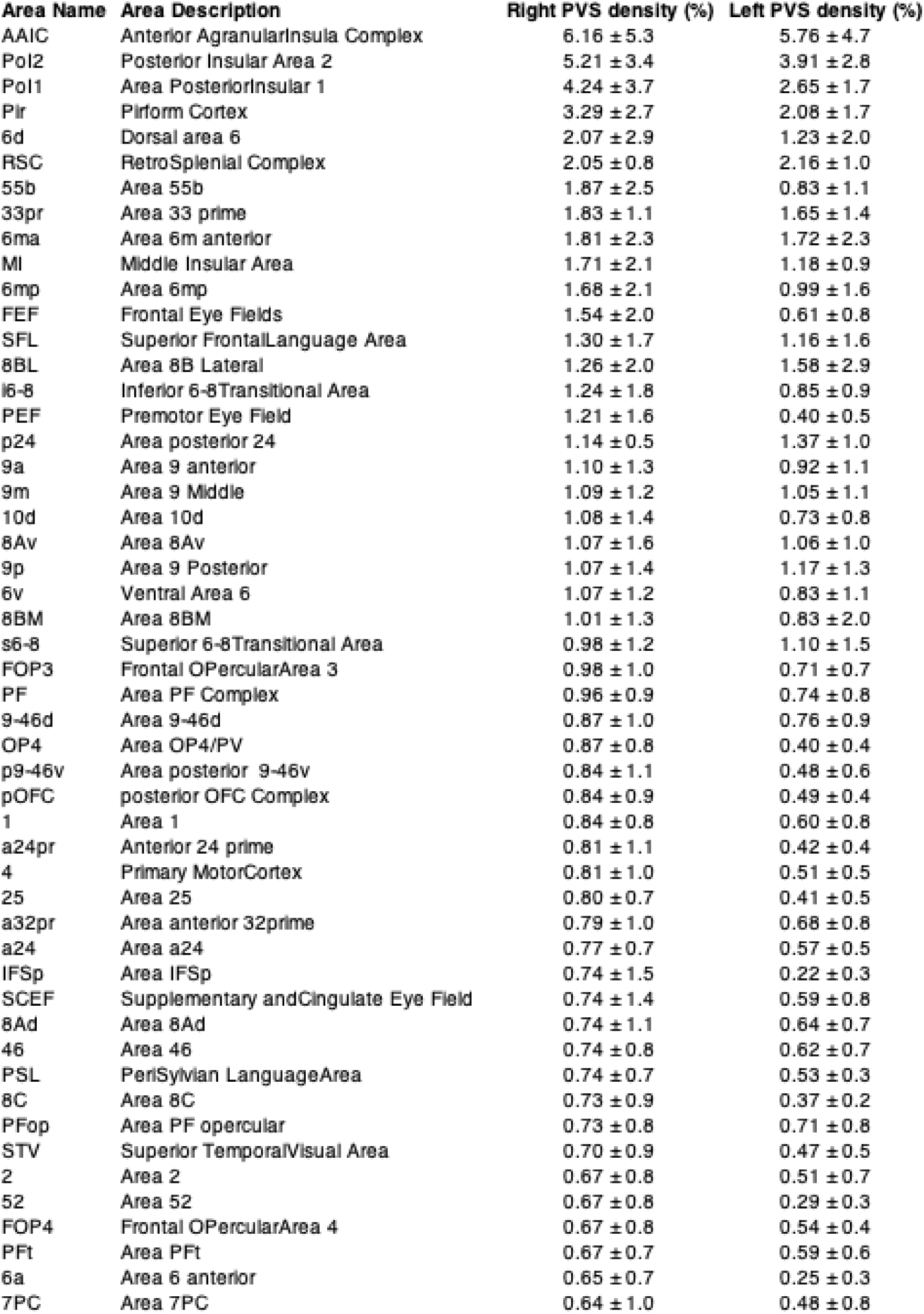
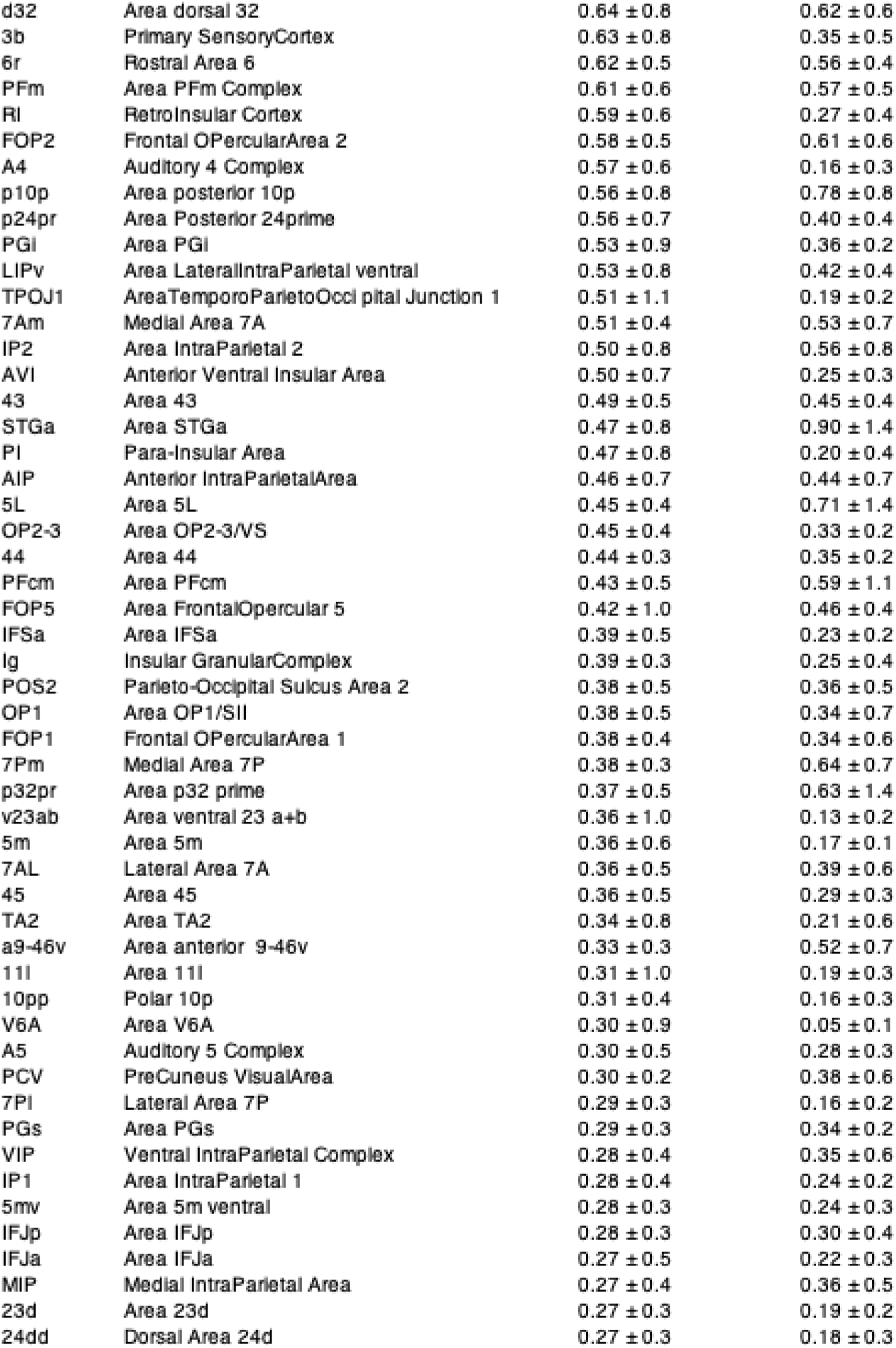
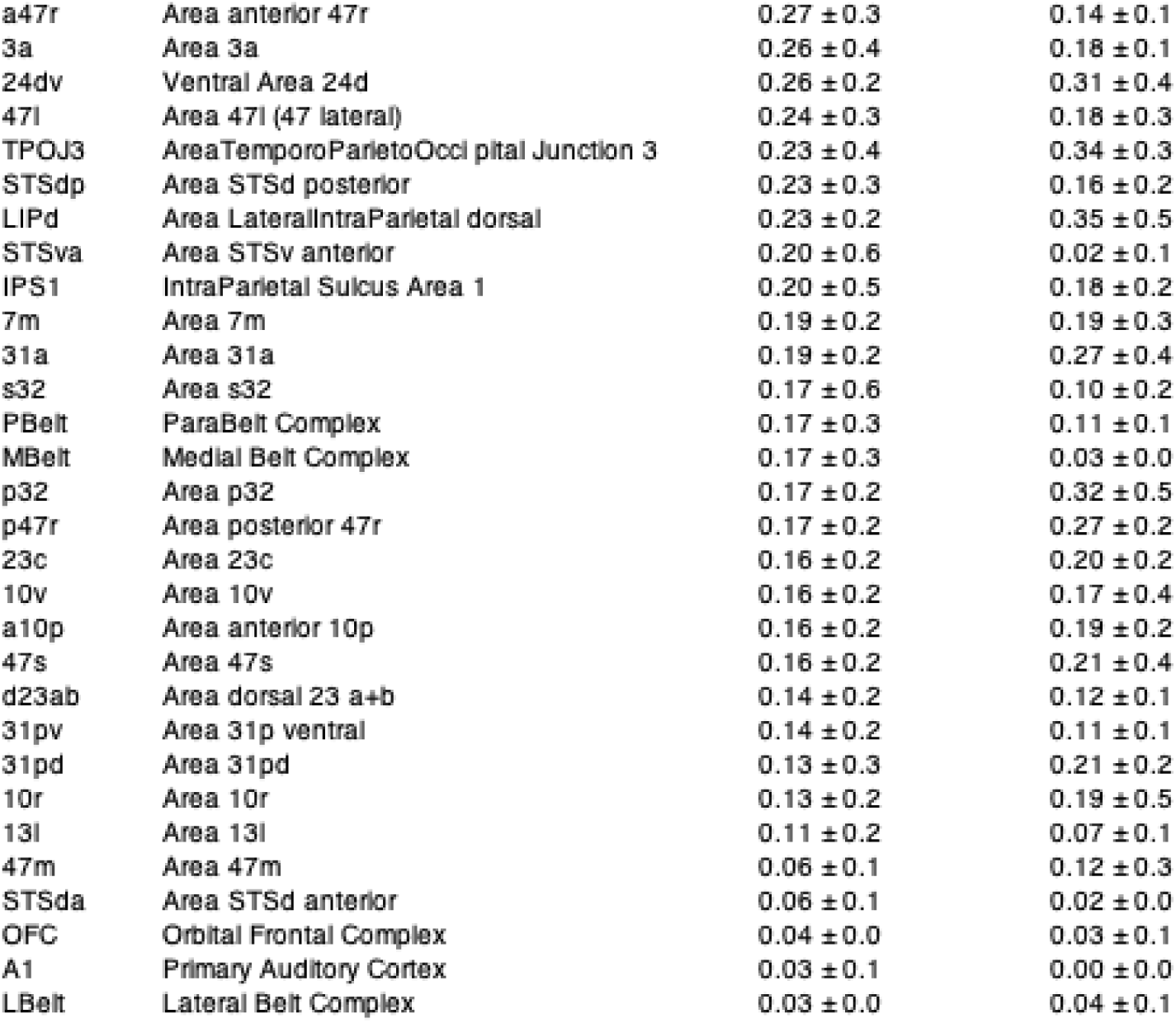
PVS densities in the 266 cortical areas for both right and left hemisphere across the 17 subjects. The cortical areas are arranged in descending order based on PVS density in the right hemisphere. The AAIC exhibits the highest density, whereas the L3belt and A1 areas show the lowest densities in the right and left hemispheres, respectively.

